# A phenome-wide association study identifies effects of copy number variation of VNTRs and multicopy genes on multiple human traits

**DOI:** 10.1101/2022.01.26.22269891

**Authors:** Paras Garg, Bharati Jadhav, William Lee, Oscar L. Rodriguez, Alejandro Martin-Trujillo, Andrew J. Sharp

**Author notes:** Address for correspondence: Andrew J. Sharp, Department of Genetics and Genomic Sciences, Icahn School of Medicine at Mount, Hess Center for Science and Medicine, 1470 Madison Avenue, Room 8-116, Box 1498, New York, NY 10029 USA.

## Abstract

The human genome contains tens of thousands of large tandem repeats and hundreds of genes that show common and highly variable copy number changes. Due to their large size and repetitive nature, these Variable Number Tandem Repeats (VNTRs) and multicopy genes are generally recalcitrant to standard genotyping approaches, and as a result this class of variation is poorly characterized. However, several recent studies have demonstrated that copy number variation of VNTRs can modify local gene expression, epigenetics and human traits, indicating that many have a functional role. Here, using read depth from whole genome sequencing to profile copy number, we report results of a phenome-wide association study (PheWAS) of VNTRs and multicopy genes in a discovery cohort of ∼35,000 samples, identifying 32 traits associated with copy number of 38 VNTRs and multicopy genes at 1% FDR. We replicated many of these signals in an independent cohort, and observed that VNTRs showing trait associations were significantly enriched for expression QTLs with nearby genes, providing strong support for our results. Fine-mapping studies indicated that in the majority (∼90%) of cases, the VNTR and multicopy genes we identified represent the causal variants underlying the observed associations. Furthermore, several lie in regions where prior SNV-based GWAS have failed to identify any significant associations with these traits. Our study indicates that copy number of VNTRs and multicopy genes contributes to diverse human traits, and suggests that complex structural variants potentially explain some of the so-called “missing heritability” of SNV-based GWAS.

## Introduction

Tandem Repeats (TRs) are stretches of DNA composed of two or more contiguous copies of a sequence of nucleotides arranged in head-to-tail pattern, e.g. CAG-CAG-CAG. The human genome contains >1 million TRs which collectively span ∼3% of our total genome.^1^ These TRs range in motif size from mono-nucleotide TRs at one extreme (e.g. TTTTTTT), to TRs with much larger motifs that can in some cases be many kilobases (kb) in size, even containing entire exons or genes within each repeated unit.^2^

Despite making a large contribution to genomic variation,^3^ historically the size and repetitive nature of TRs has meant that they are often poorly assayed or excluded from standard genotyping pipelines. This is particularly true for those repeats with moderate to large motif sizes, often termed VNTRs or macrosatellites (defined here as those with motif sizes ≥10bp). This is because the size of many VNTRs means that they generally cannot be spanned by a single Illumina sequencing read, any sequencing reads that map to multiple loci are typically removed during data processing, and microarray designs typically do not place probes within non-unique regions. As a result, unless specialized algorithms are used,^4–6^ VNTRs and other large multicopy regions are typically ignored by array or sequencing-based genome-wide association studies (GWAS).

However, numerous studies that have focused on large tandem repeats and multicopy genes have indicated their potential to influence human traits. Copy number variation (CNV) of the antimicrobial β-defensin gene cluster at 8p23.1 [MIM: 602056] has been associated with susceptibility to psoriasis,^7,8^ copy number of the Kringle repeat within *LPA* [MIM: 152200] represents the major genetic factor underlying variation in Lipoprotein A levels,^9,10^ while CNV of a 57 bp coding VNTR within *ACAN* [MIM: 155760] has a strong effect on human height.^11,12^ Similarly, CNVs of many VNTRs have been implicated as regulators of both local epigenetics and gene expression levels,^4,13–16^ and as modifiers of disease susceptibility in a variety of conditions, including Alzheimer’s,^17^ schizophrenia^18^ and ALS.^19^

Building upon our prior studies in which we assessed the regulatory potential of VNTRs,^14^ here we applied an agnostic phenome-wide association study (PheWAS) approach to determine the effect of CNV of VNTRs and multicopy genes on diverse human traits. We utilized a pipeline based on read depth for estimating copy number from short-read WGS data, and applied this to generate genotypes for 54,479 VNTRs and 1,105 copy-number variable genes that were associated with 283 traits in a discovery cohort comprising 35,254 ethnically diverse individuals sequenced as part of the NHLBI TOPMed program, followed by replication analysis in an additional cohort of 9,159 individuals. Our study identifies both known and previously unrecognized effects of CNV of large tandem repeats on a variety of human traits, and indicates broad potential for repetitive regions of the human genome to exert effects on common phenotypes.

## Materials and Methods

### Description of cohorts used for association analysis

For discovery PheWAS, we utilized WGS data from seven cohorts sequenced as part of the NHLBI TOPMed initiative (Freeze 8), accessed via dbGAP: Women’s Health Initiative (WHI) (phs001237.v2.p1) (n=11,035), Atherosclerosis Risk in Communities (ARIC) (phs001211.v4.p3) (n=3,929), Multi-Ethnic Study of Atherosclerosis (MESA) Cohort (phs001416.v2.p1) (n=5,370), The Jackson Heart Study (JHS) (phs000964.v5.p1) (n=2,777), Cardiovascular Health Study (CHS) (phs001368.v3.p2) (n=3,555), Genetic Epidemiology of COPD (COPDGene) (phs000951.v5.p5) (n=9,882), and the Framingham Heart Study (FHS) (phs000974.v4.p3) (n=4,166). For replication analysis, we utilized 10,127 individuals from The NHLBI TOPMed BioMe Biobank at Mount Sinai (phs001644.v2.p2).

Using available SNV calls, we classified each individual into one of five super-populations (European (EUR), South Asian (SAS), East Asian (EAS), African (AFR), or American (AMR)), as outlined by the gnomAD consortium.^20^ In brief, we utilized a defined set of ∼64,000 informative high quality linkage disequilibrium (LD) pruned SNVs with minor allele frequency (MAF) >5% to perform principal component analysis (PCA) using *GCTA* (v1.92.4).^21^ The results of this analysis were projected onto PCs generated from the 1000 Genomes Project cohort, and a Random Forest classifier was used to assign each sample to its major ancestry group. In doing so, we generated SNV-based PCs for each sample, which were utilized as covariates during association analysis to account for sub-structure within each super-population. In order to ensure that we only use unrelated samples in our association analysis, we generated a pairwise kinship matrix using the *KING-robust* kinship estimation in *PLINK2*.^22^ Where any samples with up to 3^rd^ degree relationships with other individuals were identified, we retained a single unrelated individual. After these QC steps, we retained data for 35,254 individuals that were used in the discovery PheWAS, and 9,159 BioMe samples that were used for replication analysis.

This study was approved by, and the procedures followed were in accordance with the ethical standards of the Institutional Review Board of the Icahn School of Medicine under HS# 19-01376.

### Phenotype data

In our discovery PheWAS, we utilized phenotype data derived from two sources: (i) 38 quantitative and binary traits that had been previously harmonized across multiple TOPMed cohorts,^23^ and (ii) data on 245 quantitative, binary, and categorical traits collected from phenotype records available within the dbGAP entries for the ARIC, CHS, JHS, MESA, and WHI cohorts. For quantitative traits, we performed quality control by removing outlier phenotype values that were ≥5 standard deviations (StDevs) from the mean for each trait. Across all 283 traits utilized, the number of individuals with data for each trait was highly variable, ranging from 370 to 33,145 (median n=5,247). For replication analyses, we utilized data for 11 quantitative traits from the Mount Sinai BioMe cohort gathered from electronic medical records for which there were data available for ≥1,000 individuals. For the 11 traits utilized, sample sizes per trait ranged from 1,534 to 8,534 (median n=8,180). In cases where an individual had repeated measurements for a trait, we utilized the median value. A complete list of the all phenotype data used is shown in Table S1.

### Targeted genotyping of VNTRs and multicopy genes

We downloaded 950,381 autosomal tandem repeats listed in the Simple Repeats track from the hg38 build of the UCSC genome browser, retaining only those repeats with motif size ≥10 base pairs (bp) and total length of repeat tract ≥100 bp. Where multiple tandem repeat annotations overlapped, these were merged together, resulting in 96,427 unique autosomal VNTR regions that were used in subsequent analysis.

In order to identify a set of genes that show frequent variation in copy number in samples of diverse ancestry, we performed *CNVnator* analysis (version 0.4.1, using default thresholds and bin size 100 bp) in 645 samples from the Human Genome Diversity Panel (HGDP) that were sequenced with PCR-free Illumina WGS.^24^ In each individual, we generated mean estimated copy number per RefSeq gene based on all coding and UTR regions, and calculated the mean and StDev of copy number per gene across the 645 samples. Doing so identified a set of 1,105 autosomal genes exhibiting putative multiple copy number states in ≥1% of samples and which showed the highest variance in copy number in the genome (henceforth referred to as multicopy genes). Many of these genes are present in multiple copies in the reference genome and/or occur in clustered gene families, are strongly over-represented within regions of segmental duplication, and have been previously identified as exhibiting high levels of CNV in the human population.^13,24,25^

In order to generate targeted copy number estimates in the TOPMed cohort, we utilized *mosdepth* on the WGS CRAMs for each TOPMed sample, to measure read depth for a user-defined set of loci.^26^ For each multicopy gene, we utilized all coding and untranslated regions per gene extended by ±100 bp and merged per gene. For VNTRs, we utilized the regions defined by Tandem Repeats Finder (as described above), and also profiled the 1 kb regions flanking each VNTR for use in quality control (described below). In addition, we also profiled a set of 200 invariant control genes for use in downstream quality control steps. Here we selected 200 highly constrained (pLI scores >0.9)^27^ autosomal genes that showed the lowest copy number variance in the HGDP. In order to correct for variations in coverage that result from fluctuations in GC-content of different genomic loci, we applied a GC-correction to the *mosdepth* output using the *DenoiseReadCounts* function in *GATK*.^28^ All normalization was based only on autosomal regions to avoid introducing technical bias between males and females resulting from differing read depths on the sex chromosomes.

To identify potential technical effects on individual samples, we generated density and PCA plots based on both the copy estimates of the 1 kb flanking regions of each VNTR and the set of 200 invariant genes, removing outlier samples. After these filtering steps, we retained a total of 34,904 samples for the analysis of multicopy genes, and 34,350 samples for the analysis of VNTRs. We calculated the StDev of each VNTR across the entire cohort, and removed VNTRs in the lowest quartile of StDev from further analysis (n=22,529). VNTRs with low sequence complexity (defined here as >80% dinucleotide repeat) were also removed (n=16,344). After filtering steps, a total of 54,479 VNTRs were used for PheWAS analysis.

In situations where a VNTR is embedded within a larger copy number variable region, copy number estimates for the VNTR based on read depth can be confounded by variations of the wider region, as these would result in gains or losses in the total number of VNTR copies present, but without any change in the length of the VNTR array. To identify VNTRs where our copy number estimates were potentially subject to this confounder, we utilized copy number analysis of 1 kb intervals at both the 3’ and 5’ regions flanking each VNTR using *mosdepth*. First, we removed any VNTR where the correlation (R) between copy number of the VNTR and both of the flanking regions was >0.5, or where both flanks were <250 bp. For the remaining VNTRs, based on the flanking regions, we identified outlier samples and filtered the VNTR genotypes of these outliers. Here, for each VNTR flank, we calculated the mean and StDev based on samples between the 30^th^ and 70^th^ percentiles of the population, defining outlier samples as those that were >7 StDevs from the mean and with consistent directionality for both 3’ and 5’ flanks. At each VNTR, genotypes for outlier samples were excluded from further analysis. Examples of this filtering step are shown in Figure S1.

Many multicopy gene families co-vary together within single CNV regions, yet are annotated with divergent names in the reference genome, e.g. the five identical copies of *PRR20* are annotated as *PRR20A* through *PRR20E* in hg38. In order to reduce redundancy when performing association testing for multicopy genes, we merged together copy number estimates for genes that fulfilled the following criteria: (i) showed highly correlated (R>0.9) *mosdepth* results across all WGS samples tested, (ii) were located on the same chromosome <1 Mb apart, and (iii) the gene name shared ≥3 letters in common. We manually inspected these merged results to ensure high specificity, and in this way generated average copy number estimates for 331 separate genes which were grouped into 104 gene families (e.g. the data for the five *PRR20* genes were summarized as a single averaged value, renamed “*PRR20_grp*”), which were utilized in subsequent PheWAS testing. Gene copy numbers aggregated in this way are shown in Table S2. After this merging step, we utilized data for a final set of 878 multicopy genes in subsequent PheWAS analysis.

As *mosdepth* only provides a read count per locus, we converted normalized *mosdepth* read counts to estimated genomic copy numbers in order to provide a more intuitive output. We utilized 225 individuals from the cohort in whom VNTRs and multicopy genes were also profiled using *CNVnator*, which provides a direct estimate of relative genomic copy number. For each locus, we used data from these 225 individuals to build a linear regression model that was used to convert the *mosdepth* output into relative genomic copy number. It should be noted that in non-unique genomic loci that contain multiple copies of a repeated motif, *CNVnator* copy number estimates represent the fold change in total (diploid) repeat number *relative to the number of motifs annotated in the (haploid) reference genome*. Thus, for the *PRR20* gene that has five copies in hg38, a *CNVnator* value of 2 corresponds to 10 copies of the gene per diploid genome.

### Phenome-wide association analysis

We performed PheWAS for discovery of associations of human traits with copy number of VNTRs and multicopy genes using *REGENIE*,^29^ incorporating covariates of sex, age, and the top three principle components derived from analysis of SNVs to account for ancestry. Additionally, we used the top five PCs derived from either invariant genes (for multicopy genes) or the 1 kb flanking regions (for VNTRs) to account for technical effects on copy number estimates from read depth. In order to minimize potential confounders such as batch effects resulting from the use of multiple different cohorts, or from traits and/or genotypes that had different frequencies among ancestries, we performed association analysis separately after dividing samples into sub-cohorts based on both TOPMed cohort (ARIC, CHS, JHS, MESA, WHI, FHS, COPD) and major ancestral group (EUR, AFR, AMR, EAS, SAS). For each association, we discarded any sub-cohorts with insufficient sample size, defined as those where phenotype data was available for <50 samples with quantitative traits, or <100 samples for binary/categorical traits. Summary results for each ancestry, and for all ancestries combined, were then generated by z-score based meta-analyses using *METAL*.^30^ We applied multiple testing corrections using both False Discovery Rate (FDR) and Bonferroni methods based on the total number of loci and traits analyzed, considering associations with <1% FDR as putatively significant. However, it should be noted that these multiple testing corrections are likely overly stringent given that some of the traits utilized are highly correlated.

For replication analysis, we selected all associations that had <1% FDR in our discovery PheWAS, and utilized data for 11 matched quantitative traits that were available in the BioMe cohort with a sample size of ≥1,000 individuals. We removed outlier phenotype values that were ≥5 StDevs from the mean for each trait, generated copy number estimates with *mosdepth*, and performed association testing using the same methodology as in the discovery PheWAS.

### Identification of causal variants using MsCAVIAR

To identify likely causal variants underlying trait associations, we utilized *MsCAVIAR*,^31^ an extension to *CAVIAR*^32^ which is suitable for the analysis of multiple cohorts of different ancestries. For each significant VNTR or multicopy gene identified in the discovery PheWAS, we identified all SNVs with MAF ≥1% located within ±100 kb, excluding those that failed tests for Hardy-Weinberg equilibrium (p<10^−300^), had genotyping rate <90%, or had <5 observations of the minor allele in any of the sub-cohorts analyzed. We then performed association analysis of each SNV with the trait using *REGENIE* followed by meta-analysis using *METAL*, utilizing the same methodology and covariates as used in the original PheWAS. Pairwise correlation was used to define the LD structure for SNVs in each cohort/ancestry. The top 100 most significantly associated SNVs at each locus, along with genotype of the respective VNTR/multicopy gene, were then used as input into *MsCAVIAR* together with the VNTR/multicopy gene to infer the most likely causal variant at each locus using default thresholds with rho probability (-r) of 0.95, gamma (-g) of 0.01, and using two as the maximum number of causal variants.

### Association analysis of multicopy genes with local gene expression

As the original GTEx analysis included a requirement for RNAseq reads to have unique alignments within the reference genome in order to be considered, we hypothesized that there would be systematic bias in the measurement of expression levels for many multicopy genes, e.g. in GTEx data the five *PRR20* genes are listed as being not expressed across all human tissues, likely due to RNAseq reads for these genes having non-unique mapping and thus being discarded. To identify potential effects of CNV of multicopy genes on their own expression level, we therefore performed a re-analysis of GTEx data in which we omitted the requirement for RNAseq reads to have unique alignments within the reference genome. We utilized Illumina WGS data for 394 samples from the GTEx project that had been sequenced using PCR-free WGS. We downloaded RNAseq data for these samples, and performed a re-analysis using an identical pipeline to the GTEx consortium, except for the use of featureCounts that allowed multi-mapping reads to be included in the analysis (with parameters set to -t exon -g gene_id --minOverlap 20 --fracOverlap 0.5 -M -O -Q 0 --ignoreDup). We considered associations with <10% FDR as putatively significant.

For the *PRSS1/PRSS2* locus, we also performed a *cis* eQTL analysis using data from 676 individuals from the PPMI cohort for which both RNAseq data derived from whole blood and WGS data were available. We used copy number estimates for *PRSS1/PRSS2* derived from *CNVnator* analysis, Gencode gene annotations, and utilizing a methodology as described previously.^14^

## Results

### Copy number estimates of VNTRs and multicopy genes in ∼45,***000 individuals***

Using normalized read depth from Illumina WGS data as a proxy for diploid copy number, we profiled ∼45,000 samples sequenced as part of the TOPMed program, generating copy number estimates for a set of 54,479 VNTRs and for a set of 1,105 autosomal genes showing common CNV in a diverse human population.

Example data for the □-defensin gene cluster is shown in Figure 1A, and for salivary amylase gene cluster in Figure S2. Given the number of individuals we profiled was much larger than in previous studies,^13,24,25^ we observed instances of rare individuals exhibiting extremes of copy number, with estimated copy numbers for some genes that were 10-20 times greater than the population average. Examples of genes showing highly amplified copy number include *HPR* [MIM: 140210], *CCL3L1/CCL4L1/TBC1D3B* [MIM: 601395/603782/610144], the salivary amylases *AMY1* [MIM: 104700/104701/104702], protocadherins *PCDHB7/PCDHB8* [MIM: 606333/606334], α-defensins *DEFA* [MIM: 125220/604522], and *ORM1/ORM2* [MIM: 138600/138610]. (Figure 1B, Figure S3). At the other extreme, we also observed one individual who apparently completely lacked the entire □-defensin gene cluster at 8p23.1 (Figure S4).

**Figure 1.**
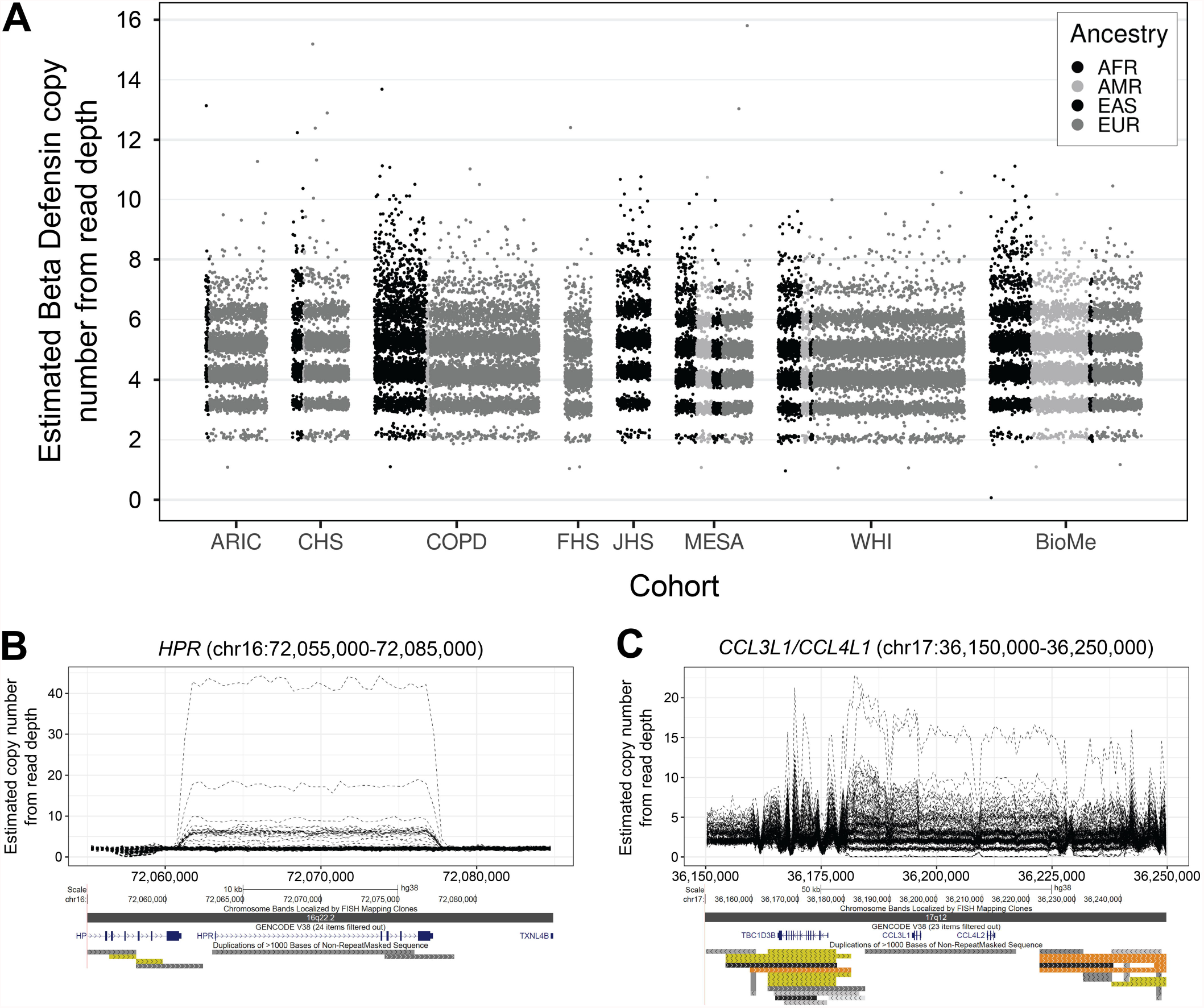
Identification of extreme variations in gene copy number in the human population. **(A)** Diploid copy number estimates generated using *mosdepth* for the □-defensin gene family at 8p23.1 in ∼45,000 individuals from eight TOPMed cohorts used in this study. While most individuals carry between 2-8 copies of the □-defensin locus, we observed rare individuals with up to 16 copies, and conversely one individual who apparently completely lacked □-defensin (Figure S4). Other examples of genes exhibiting extreme variations in copy number include **(B)** *HPR*, where one individual carried an estimated ∼42 copies, compared to a median of two in the general population, and **(C)** *CCL3L1/CCL4L1*. Plots in **(B)** and **(C)** show *CNVnator* relative diploid copy number per 500 bp bin in 225 selected individuals. Below each plot is an image of the region taken from the UCSC Genome Browser showing gene and segmental duplication annotations. Additional examples are shown in Figure S3.

We compared our copy number estimates derived from read depth to those generated by other approaches. Previously, using a set of samples sequenced with both Pacific Biosciences long reads and Illumina WGS, we had observed a high correlation (R^2^=0.81) between VNTR copy estimates derived from read depth with those obtained via long-read sequencing.^14^ A previous study had utilized the paralog ratio test (PRT) to estimate □-defensin copy number in ∼1,000 Europeans.^7^ Although we were unable to genotype these same samples, based on read depth, we observed an almost identical distribution of copy number estimates for L-defensin in TOPMed samples of European ancestry compared to these samples previously genotyped using the PRT, suggesting that the two methods yield highly concordant results (Figure S5). Similar results were also obtained when comparing copy number estimates for salivary amylase generated by read depth with previously published data generated using quantitative PCR (qPCR) (Figure S2).

### Phenome-wide association analysis of VNTRs and multicopy genes

To assess the potential influence of copy number changes of VNTRs and multicopy genes on human traits, we performed PheWAS in a discovery cohort comprising ∼35,000 individuals of diverse ancestry derived from seven different cohorts that had been sequenced as part of the NHLBI TOPMed study. Considering results based on the meta-analyses of all ancestries combined, at a threshold of <1% FDR, we identified 21 pairwise associations with VNTRs (Table S3) and 42 pairwise trait associations with multicopy genes (Table S4). Using QQ-plots to explore the distribution of observed versus expected associations, we observed a clear enrichment for significant associations compared to the null distribution, with little evidence of genomic inflation for either VNTRs or multicopy genes (values of λ=1.02 and 1.04, respectively) (Figure S6). The genome-wide distribution of association signals for VNTRs and multicopy genes are shown in Figure 2.

**Figure 2.**
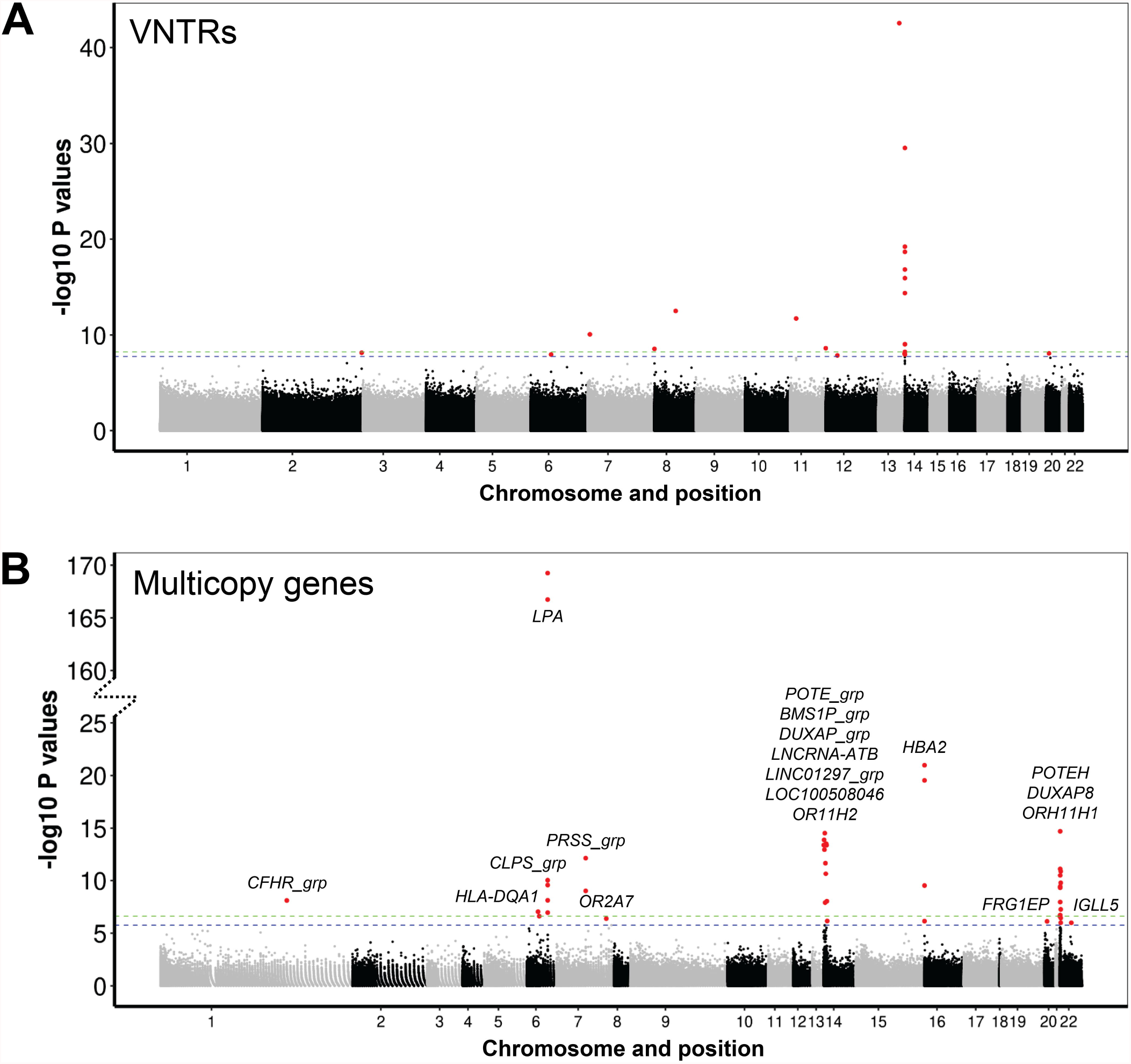
Manhattan plots showing genomic location of variants identified in a PheWAS using 283 traits and. **(A) 54**,**479 VNTRs and (B) 878 multicopy genes in ∼35**,**000 TOPMed individuals**. Name(s) of multicopy genes are shown adjacent to each significant association in the lower panel. Note the discontinuous y-axis used to display the results for multicopy genes, resulting from the very strong association of *LPA* copy number with Lipoprotein A levels. Dashed green and blue horizontal lines indicate the p=0.05 Bonferroni and 1% FDR significance thresholds, respectively. Points in red indicate significant associations at <1% FDR. Full results are shown in Tables S3 and S4.

We identified several significant associations that have been previously reported in the literature, indicative that our methodology detected robust associations. These included an inverse association of *LPA* copy number with Lipoprotein A levels,^9,10^ and reduced *HBA2* copy number associated with multiple red blood cell traits [MIM: 141850].^33^ Also notable was a previously reported association of a coding VNTR in *ACAN* with standing body height,^11,12^ which in our analysis achieved FDR q=0.011, thus just missing our statistical significance threshold. In addition, our PheWAS also identified *LPA* copy number as influencing risk of percutaneous transluminal coronary angioplasty, a common surgical intervention for cardiovascular disease, indicating we were able to detect phenotypic consequences downstream of the primary influence of *LPA* copy number. Other biologically plausible associations identified include a VNTR located within intron 2 of *F7* (*Factor VII*) [MIM: 613878] showing strong association with circulating factor VII levels, and a VNTR located within intron 1 of *FADS2* (*Fatty Acid DeSaturase 2*) [MIM: 606149] associated with levels of specific fatty acids.

We also identified multiple VNTRs located at 14q11.2 that were associated with levels of interleukin-6, fibrinogen and multiple immune cells in the blood. Notably, these VNTRs are all embedded within a large cluster of T cell receptor α genes (*TRA*), and three of them had been identified in prior analyses as expression quantitative trait loci (eQTLs) for nearby *TRA* genes. Overall, comparing results with our previous study that identified VNTRs that act as expression QTLs in the GTEx and PPMI cohorts,^14^ we observed that VNTRs showing significant trait associations were significantly enriched for eQTLs when compared to the background set of all VNTRs tested (5.2-fold enrichment, p=5.4×10^−4^).

### The influence of gene copy number on local gene expression

In previous work, we identified VNTRs that act as eQTLs and methylation QTLs.^14^ Here, by performing a re-analysis of WGS and RNAseq data from 48 tissues released by the GTEx consortium, we extended this approach to also identify associations between gene copy number and (i) their own expression level, or (ii) the expression level of other nearby genes. In total, at a 10% FDR threshold, we identified 217 multicopy genes that were significantly associated with their own copy number in one or more tissues. Of these, 98.1% of the pairwise correlations showed positive directionality, indicating that for many multicopy genes, increased genomic copy number results in increased gene expression, as expected (Table S5).

However, in some cases we also identified more complex effects, such as examples where neighboring copy number invariant genes also showed altered expression levels. Representative examples of the patterns observed at the *RHD* and *HPR* loci are shown in Figure 3. While the expression of both *RHD* and *HPR* show strong positive correlations with their own copy number across nearly all tissues analyzed, at both loci the expression level of multiple neighboring genes are also correlated with copy number. At the *HPR* locus, we identified six neighboring genes within ±100 kb that all showed positive correlations with *HPR* copy number, despite all six genes being located outside the *HPR* CNV region. At the *RHD* locus, while three genes that lie within the common CNV region were all strongly positively associated with *RHD* copy number, three other genes that lie outside of the CNV also showed expression levels that correlated variably with *RHD* copy number, including one gene (*TMEM50A*) whose expression was positively correlated with *RHD* copy number in some tissues, and negatively correlated with *RHD* copy number in other tissues. These results indicate that many CNVs have wider effects that can often alter the regulation of other genes *in cis*.

**Figure 3.**
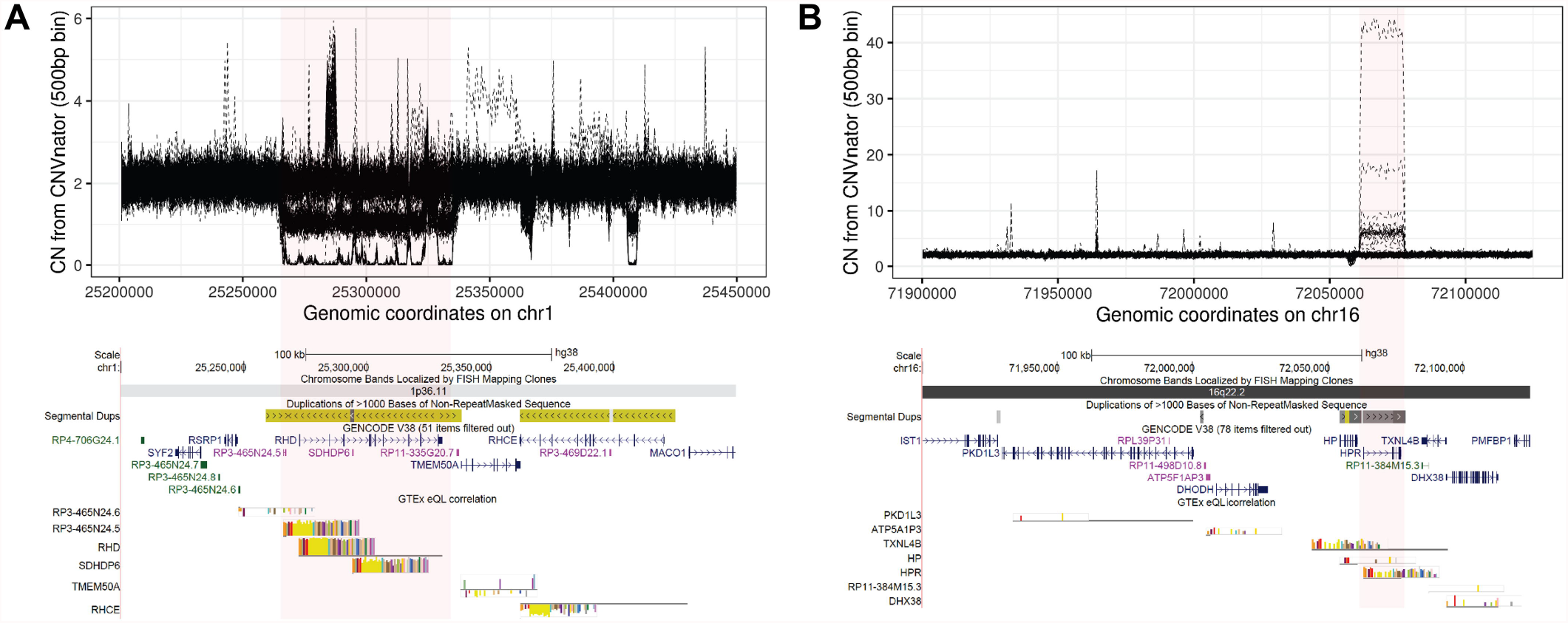
Genomic copy number of multicopy genes often correlates with their own expression level and that of multiple neighboring genes *in cis*. **(A)** The *RHD* locus (chr1:25,200,000-25,450,000) and **(B)** the *HPR* locus (chr16:71,900,000-72,125,000). At the base of each plot, the colored bar plots show custom UCSC Genome Browser tracks indicating significant (<10% FDR) correlation (R) values between estimated copy number of the CNV (red shaded) region and gene expression level across the 48 GTEx tissues analyzed. Direct correlations are indicated by positive R values (i.e. projecting up above the baseline), while inverse correlations are indicated by negative R values (i.e. projecting down below the baseline). For both *RHD* and *HPR*, increased genomic copy number resulted in increased expression (i.e. positive correlations) for genes within the CNV region. In addition, despite being located outside the CNV region, the expression level of multiple other neighboring genes also showed either positive and/or negative correlations with copy number of *RHD* and *HPR*. The upper plot in each panel shows *CNVnator* relative diploid copy number per 500 bp bin in 225 selected individuals, with the common copy number variable region shaded in red. The lower plot in each panel shows an image of the region taken from the UCSC Genome Browser showing gene and segmental duplication annotations, in addition to significant eQTL results. Complete eQTL data for all multicopy genes in the GTEx cohort are shown in Table S5.

One notable example of this phenomenon was the *PRSS1*/*PRSS2* locus [MIM: 276000/601564], copy number of which was associated with circulating lymphocyte and white blood cell levels. Our eQTL analysis identified that *PRSS1*/*PRSS2* copy number was associated with the expression level of multiple neighboring *TRB* (T cell receptor □) genes, suggesting a potential link to explain how CNV of a digestive enzyme could lead to altered immune function (Figure S7).

### Replication in an independent cohort

In order to assess the robustness of the associations we identified in the discovery cohort, we conducted replication analysis using 9,159 individuals from the BioMe cohort for which both WGS and phenotype data were available. Here, we focused on 11 quantitative traits that showed significant associations (<1% FDR) with either VNTRs and/or multicopy genes in our discovery cohort, and for which data were available in at least 1,000 BioMe individuals. Utilizing an identical methodology to the discovery PheWAS, six of the eight VNTR:trait pairs tested showed strong replication in BioMe, with replication p-values for these associations ranging from 2.4×10^−25^ to 5.2×10^−51^ (Table S6). Similarly, nine of 17 multicopy gene:trait pairs tested showed replication in at least one ancestry in BioMe, with replication p-values ranging from 0.04 to 1.6×10^−133^ (Table S7).

### Fine mapping indicates most VNTR and multicopy gene variations associations are causal

Due to the LD structure of the genome, multiple different genetic variants within a locus may show association with a trait, but only one or a few of these are likely to be causal. In order to assess whether the VNTRs and multicopy genes identified in our PheWAS represent the likely causal variants responsible for the observed association signals, we applied *MsCAVIAR*.^31^ Using this approach, we observed that for 24 of 28 (86%) VNTR:trait associations and 40 of 42 (95%) multicopy gene:trait associations, the VNTR/multicopy gene was ranked as the most likely causal variant compared to local SNVs (Tables S8 and S9).

Closer examination of some of these regions corroborated this result. Figure 4 shows the example of a 37mer VNTR located within a large cluster of T cell receptor genes at 14q11.2 (chr14:22,355,658-22,355,834) that was scored by *MsCAVIAR* as the likely causal variant at this locus associated with lymphocyte concentration in blood. In our discovery PheWAS, we identified a strong and consistent association between copy number of this VNTR and lymphocyte concentration across all TOPMed cohorts and ancestries tested (discovery meta-analysis p=2.9×10^−30^) and which strongly replicated in the BioMe cohort (p=5.2×10^−51^). Our previous association analysis in the PPMI cohort had identified this same VNTR as an eQTL of the nearby transcript ENSG00000256221.1,^14^ supporting this as a potentially functional variant. However, no prior GWAS have reported any association signals for lymphocyte concentration in this region.^34^ We confirmed the absence of local associations between lymphocyte concentration and local SNVs by repeating the association analysis using all SNVs located within ±100 kb of the VNTR, which identified no significant signals. Applying *MsCAVIAR*, we confirmed that copy number of the VNTR was the most likely causal variant to explain the observed association with lymphocyte concentration (posterior p=0.988) (Table S8). Of note, the same VNTR was also significantly associated with white cell count, neutrophil count, and interleukin 6 levels.

**Figure 4.**
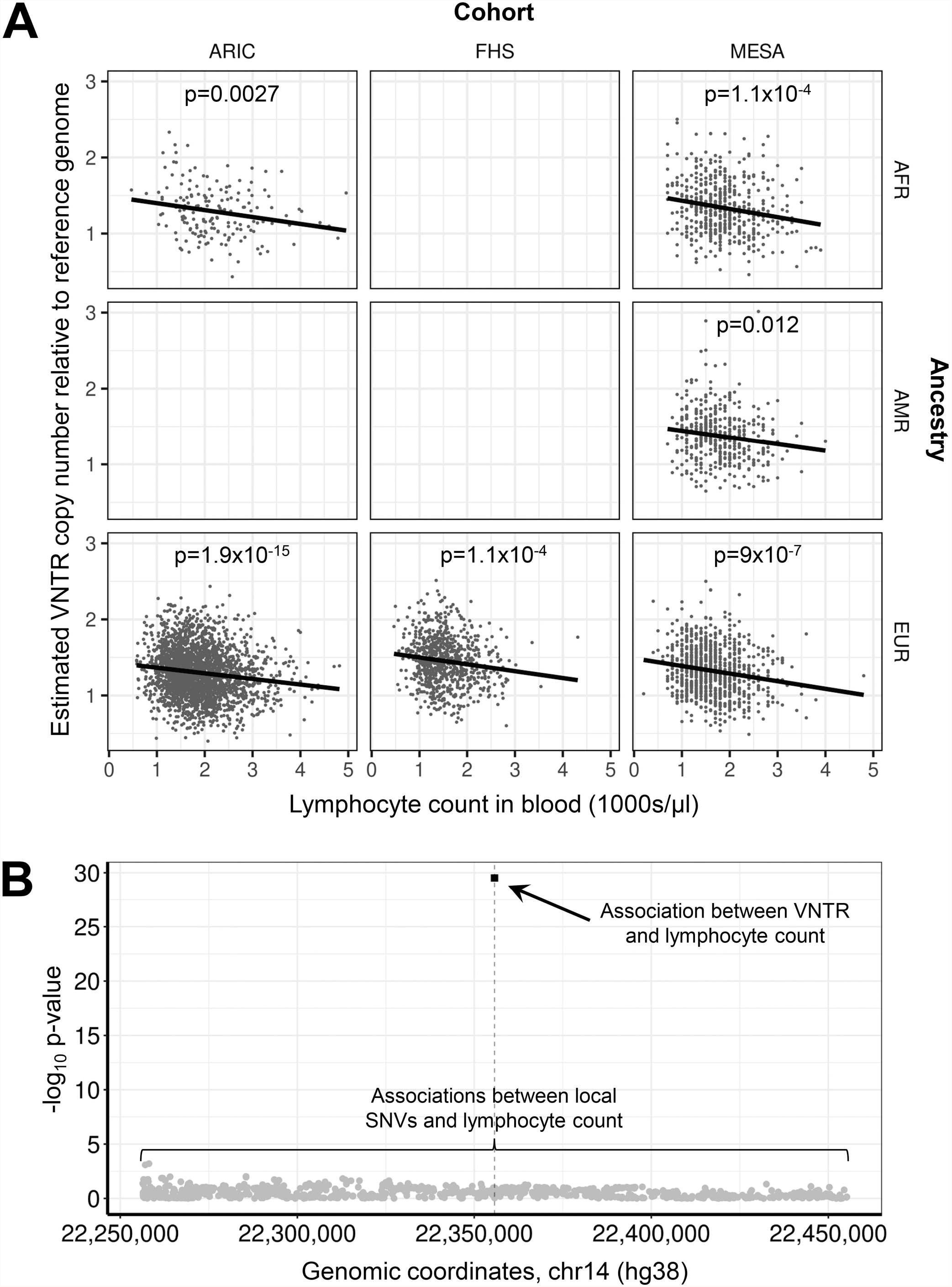
Copy number of a 37mer VNTR located within a large cluster of T cell receptor genes at 14q11.2 (chr14:22,355,658-22,355,834) is the likely causal variant associated with lymphocyte concentration in blood. **(A)** We identified a strong and consistent association between copy number of this VNTR and lymphocyte concentration across all TOPMed cohorts and ancestries tested (discovery meta-analysis p=2.9×10^−30^). In contrast, no prior GWAS have reported signals for lymphocyte concentration in this region.^34^ **(B)** We confirmed the absence of SNV associations in this region by repeating the association analysis with lymphocyte concentration using all SNVs located within ±100 kb of the VNTR (grey circles), which all yielded non-significant p-values compared to the VNTR (black square). *MsCAVIAR* confirmed the VNTR as the single most likely causal variant to explain the observed association with lymphocyte concentration (posterior p=0.988) (Table S8). The same VNTR was also significantly associated with white cell count, neutrophil count, and interleukin 6 levels.

In contrast, we performed a similar analysis of a 34mer tandem motif located within intron 1 of the *F7* gene at 13q34 (chr13:113,107,242-113,109,277) (Figure S8). Although this VNTR showed a strong and consistent association with Factor VII levels in blood across multiple TOPMed cohorts (discovery meta-analysis p=2.85×10^−43^), multiple prior GWAS have reported associations for Factor VII levels in this region.^34^ Consistent with this, *MsCAVIAR* ranked 24 nearby SNVs each as having a higher probability of being potentially causal compared to the VNTR. We confirmed this by repeating the association analysis using all SNVs located within ±100 kb of the VNTR, identifying >20 local SNVs which showed stronger association with Factor VII levels than observed for VNTR copy number, consistent with the notion that the VNTR is unlikely to be the causal variant at this locus.

## Discussion

Almost all published GWAS have utilized genotyping data from SNVs, which are typically assayed using microarrays. One of the fundamental assumptions underlying SNV-based GWAS is that due to the LD architecture of the human genome, the true causal variants that drive association signals at a locus do not have to be directly genotyped, as long as these causal variants can be “tagged” by common flanking SNVs. However, a resulting limitation of this approach is that any genomic variant that is neither directly genotyped, nor shows sufficient LD with flanking SNVs, is not effectively assayed, and therefore remains essentially invisible to standard SNV-based GWAS.

Here, we utilized an alternate approach based on read depth to specifically genotype large polymorphic tandem repeats that are typically ignored by most genomic studies. Using these as input for a PheWAS, we were able to identify several dozen VNTRs and multicopy genes, CNV of which are associated with a variety of human traits. This included several associations that have been previously reported in the literature, including the Kringle repeat of *LPA* copy that influences Lipoprotein A levels,^9,10^ a coding VNTR within *ACAN* that has a strong effect on height,^11,12^ and reduced *HBA2* copy number that influences multiple red blood cell traits,^33^ thus validating our methodology. Furthermore, we were able to replicate many signals in a second cohort.

Due to the LD architecture of the genome that often results in multiple variants within a region that are all associated with a trait, a fundamental question in the field of association analysis is the identification of causal variants, i.e. those that are primarily responsible for the observed association, rather than simply being “passengers” that are in LD with the true causal variant. As we primarily investigated VNTRs and multicopy genes, to assess whether these represented the causal variants underlying the association signals identified, we applied fine-mapping approaches to compare them against local SNVs. This analysis indicated that the large majority of associations we identified (86-95%) are likely causal for the associations we report. At several of these loci, no prior associations with the trait had been identified by SNV-based GWAS, suggesting that changes in copy number of these VNTRs are not sufficiently tagged by flanking SNVs to be detectable in standard GWAS. Thus, by specifically focusing on VNTRs and multicopy genes, i.e. complex variants that are typically ignored in most genomic studies, we were able to identify novel causal variants contributing to human phenotypic variation that seemingly remain invisible to standard GWAS approaches that rely on genotyping SNVs. As such, consistent with prior hypotheses, our data provides strong support that complex structural variants underlie at least some of the so-called “missing heritability” of GWAS.^35^

Given the relatively large sample size we used in comparison to prior studies, we were able to identify several loci exhibiting much more extreme copy number changes in the human population than have been previously reported. For example, a prior study of the Human Genome Diversity Panel reported that *HPR* varies from 2-9 copies in the global population,^24^ with the highest copy number observed in African individuals, where increased copy number is thought to be confer resistance to trypanosome infection.^36^ However, we identified individuals carrying up to an estimated 42 copies of this gene, and observed similar extreme copy numbers for several other genes. Such focal amplifications likely occur through a mechanism of non-allelic homologous recombination that generate long tandem arrays, and have been reported previously at loci such as □-defensin and *REXO1L1*, where extreme copy number amplifications can occasionally lead to cytogenetically visible euchromatic variants that can span multiple megabases.^37,38^

Although the overwhelming majority of VNTRs we tested are non-coding, we observed a significant enrichment for those associated with traits by PheWAS to act as eQTLs for nearby genes, indicating that in many cases copy number changes of VNTRs likely exert their effects on human phenotypes through altered regulation of local gene expression. In the case of multicopy genes, by definition, CNV of these regions results in copy number changes to entire gene(s). As such, the most parsimonious explanation of how CNV of a gene leads to an altered phenotype is that changes in gene copy number result in concomitant alterations in the expression level of the gene (and presumably therefore the resulting protein level). We specifically investigated this hypothesis by performing an eQTL study in which we first re-analyzed GTEx data, but allowed RNAseq reads with multiple genomic alignments to be considered when quantifying gene expression levels, a methodology which is likely better suited than the original GTEx pipeline for characterizing genes that often have multiple copies in the reference genome. Indeed, consistent with the expectation that increased genomic copy number of a gene will result in increased expression, the overwhelming majority (>98%) of significant correlations showed positive directionality. However, somewhat surprisingly, we also observed that in many cases, CNV of some multicopy genes also results in altered expression of other neighboring genes *in cis*, even where copy number of these neighboring gene remained constant. This has also been noted previously in studies of much larger recurrent microdeletion/duplication regions^39–41^ and thus seems to be a common effect of many copy number variants. Notably, this phenomenon provides a biologically plausible mechanism to explain how CNV of *PRSS1*/*PRSS2*, which represent the major digestive trypsinogens secreted by the pancreas into the gut, could influence the level of circulating immune cells through regulatory effects on the expression of nearby T cell receptor □ genes.

While the analysis approach we used here has certain advantages, the use of read depth for typing VNTRs also has several limitations, as follows: (i) Read depth does not provide any allelic information, and only yields a relative estimate of total copy number from the sum of both alleles; (ii) It is unable to differentiate between divergent repeat motifs that may independently vary in copy number; (iii) As mapping of reads to a VNTR is based on alignment to the reference sequence, repeat motifs that diverge from the reference genome would likely be poorly assayed or missed entirely; (iv) The ability of read depth for genotyping VNTRs is inversely related to both motif size and copy number, and thus copy number estimates of VNTRs with shorter motifs and lower copy number will tend to have lower accuracy; (v) The use of read depth can also be confounded through batch effects in WGS data, or where a VNTR is contained within a larger region of CNV. However, we applied stringent quality control steps to remove such confounders (Figure S1 and Methods), and performed replication and eQTL studies in separate cohorts to provide secondary support for many of the associations we identified.

Although we also performed separate analyses of each major human ancestry, the resulting reduction in sample size meant that we had very limited power to identify ancestry-specific associations. The only clear example of this that we were able to identify was the association of reduced *HBA2* copy number with multiple red blood cell traits, all of which were highly significant in individuals of African ancestry, but barely reached nominal significance in Europeans, despite having a much larger sample size. This observation is consistent with the known prevalence of □-thalassemia in regions where malaria is endemic, where heterozygous deletions of *HBA2* are thought to provide enhanced immunity against malaria.^42^

Finally, it should also be noted that although we utilized data for ∼33,000 individuals in our discovery PheWAS, phenotype data for most of the traits tested in our discovery cohort were available only for a fraction of these, with a median of 5,247 phenotyped individuals per trait. Given this relatively modest sample size for the majority of traits we studied, statistical power was a limiting factor in this study. We predict that the use of larger cohorts will reveal much more extensive effects of VNTRs and multicopy on diverse human traits and has the potential to uncover genetic predispositions that are recalcitrant to standard SNV-based GWAS.

## Data Availability

All data produced in the present study are available upon reasonable request to the authors. Datasets used in this study are available as follows:
Database of Genotypes and Phenotypes (dbGaP), NHLBI TOPMed: Women's Health Initiative (WHI) https://www.ncbi.nlm.nih.gov/projects/gap/cgi-bin/study.cgi?study_id=phs001237.v2.p1
Database of Genotypes and Phenotypes (dbGaP), NHLBI TOPMed - NHGRI CCDG: Atherosclerosis Risk in Communities (ARIC), https://www.ncbi.nlm.nih.gov/projects/gap/cgi-bin/study.cgi?study_id=phs001211.v4.p3
Database of Genotypes and Phenotypes (dbGaP), NHLBI TOPMed: MESA and MESA Family AA-CAC, https://www.ncbi.nlm.nih.gov/projects/gap/cgi-bin/study.cgi?study_id=phs001416.v2.p1 
Database of Genotypes and Phenotypes (dbGaP), NHLBI TOPMed: The Jackson Heart Study (JHS), https://www.ncbi.nlm.nih.gov/projects/gap/cgi-bin/study.cgi?study_id=phs000964.v5.p1 
Database of Genotypes and Phenotypes (dbGaP), NHLBI TOPMed: Trans-Omics for Precision Medicine (TOPMed) Whole Genome Sequencing Project: Cardiovascular Health Study, https://www.ncbi.nlm.nih.gov/projects/gap/cgi-bin/study.cgi?study_id=phs001368.v3.p2 
Database of Genotypes and Phenotypes (dbGaP), NHLBI TOPMed: Genetic Epidemiology of COPD (COPDGene), https://www.ncbi.nlm.nih.gov/projects/gap/cgi-bin/study.cgi?study_id=phs000951.v5.p5 
Database of Genotypes and Phenotypes (dbGaP), NHLBI TOPMed: Genomic Activities such as Whole Genome Sequencing and Related Phenotypes in the Framingham Heart Study, https://www.ncbi.nlm.nih.gov/projects/gap/cgi-bin/study.cgi?study_id=phs000974.v4.p3 
Database of Genotypes and Phenotypes (dbGaP), NHLBI TOPMed - NHGRI CCDG: The BioMe Biobank at Mount Sinai, https://www.ncbi.nlm.nih.gov/projects/gap/cgi-bin/study.cgi?study_id=phs001644.v2.p2 
Database of Genotypes and Phenotypes (dbGaP), GTEx data, https://www.ncbi.nlm.nih.gov/projects/gap/cgi-bin/study.cgi?study_id=phs000424.v7.p2 
GTEX portal, https://www.gtexportal.org/
Human Genome Diversity Panel, https://www.internationalgenome.org/data-portal/data-collection/hgdp
Parkinsons Progression Markers Initiative (PPMI), https://www.ppmi-info.org/

## Description of Supplemental Data

Supplemental Data include eight Supplemental Figures, and nine Supplemental Tables.

## Acknowledgements

This work was supported by NIH grants NS105781 and NS120241 to AJS, NIH predoctoral fellowship NS108797 to OR, and NHLBI BioData Catalyst Fellowship 5120339 to AMT. Research reported in this paper was supported by the Office of Research Infrastructure of the National Institutes of Health under award number S10OD018522. The content is solely the responsibility of the authors and does not necessarily represent the official views of the National Institutes of Health. This work was supported in part through the computational resources and staff expertise provided by Scientific Computing at the Icahn School of Medicine at Mount Sinai. A full list of acknowledgements for all datasets used in this study are listed in the Supplemental Data file.

## Data and Code Availability

Database of Genotypes and Phenotypes (dbGaP), NHLBI TOPMed: Women’s Health Initiative (WHI) https://www.ncbi.nlm.nih.gov/projects/gap/cgi-bin/study.cgi?study_id=phs001237.v2.p1

Database of Genotypes and Phenotypes (dbGaP), NHLBI TOPMed - NHGRI CCDG: Atherosclerosis Risk in Communities (ARIC), https://www.ncbi.nlm.nih.gov/projects/gap/cgi-bin/study.cgi?study_id=phs001211.v4.p3

Database of Genotypes and Phenotypes (dbGaP), NHLBI TOPMed: MESA and MESA Family AA-CAC, https://www.ncbi.nlm.nih.gov/projects/gap/cgi-bin/study.cgi?study_id=phs001416.v2.p1

Database of Genotypes and Phenotypes (dbGaP), NHLBI TOPMed: The Jackson Heart Study (JHS), https://www.ncbi.nlm.nih.gov/projects/gap/cgi-bin/study.cgi?study_id=phs000964.v5.p1

Database of Genotypes and Phenotypes (dbGaP), NHLBI TOPMed: Trans-Omics for Precision Medicine (TOPMed) Whole Genome Sequencing Project: Cardiovascular Health Study, https://www.ncbi.nlm.nih.gov/projects/gap/cgi-bin/study.cgi?study_id=phs001368.v3.p2

Database of Genotypes and Phenotypes (dbGaP), NHLBI TOPMed: Genetic Epidemiology of COPD (COPDGene), https://www.ncbi.nlm.nih.gov/projects/gap/cgi-bin/study.cgi?study_id=phs000951.v5.p5

Database of Genotypes and Phenotypes (dbGaP), NHLBI TOPMed: Genomic Activities such as Whole Genome Sequencing and Related Phenotypes in the Framingham Heart Study, https://www.ncbi.nlm.nih.gov/projects/gap/cgi-bin/study.cgi?study_id=phs000974.v4.p3

Database of Genotypes and Phenotypes (dbGaP), NHLBI TOPMed - NHGRI CCDG: The BioMe Biobank at Mount Sinai, https://www.ncbi.nlm.nih.gov/projects/gap/cgi-bin/study.cgi?study_id=phs001644.v2.p2

Database of Genotypes and Phenotypes (dbGaP), GTEx data, https://www.ncbi.nlm.nih.gov/projects/gap/cgi-bin/study.cgi?study_id=phs000424.v7.p2

GTEX portal, https://www.gtexportal.org/

Human Genome Diversity Panel, https://www.internationalgenome.org/data-portal/data-collection/hgdp

Parkinson’s Progression Markers Initiative (PPMI), https://www.ppmi-info.org/

## Web Resources

GWAS catalog, https://www.ebi.ac.uk/gwas/

OMIM, http://www.omim.org/

UCSC Genome Browser, http://genome.ucsc.edu

