## Supplemental Figures for "A phenome-wide association study identifies effects of copy number variation of VNTRs and multicopy genes on multiple human traits"

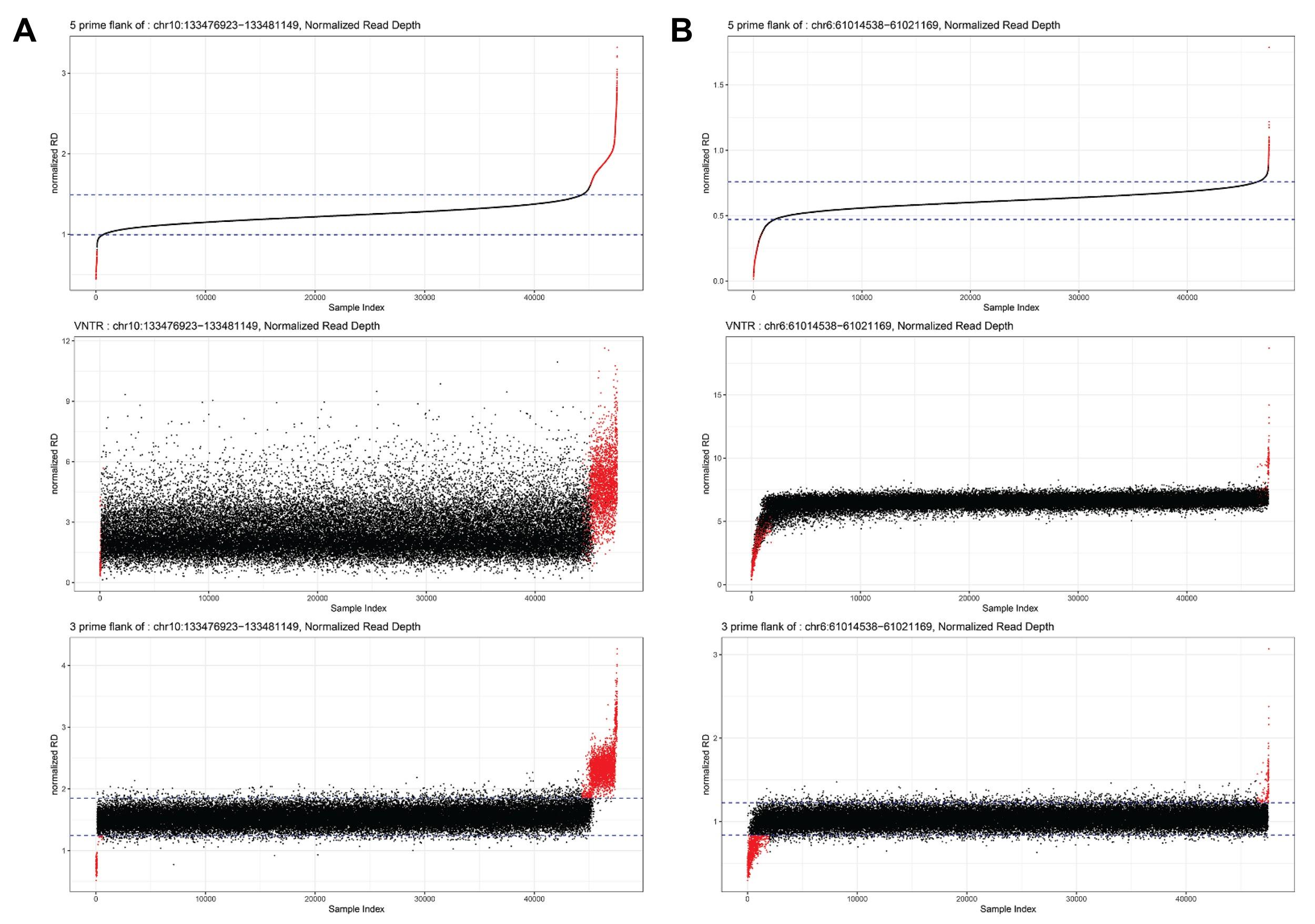

**Figure S1. Use of copy number estimates for VNTR flanks to remove outlier samples where VNTR estimates are likely erroneous due to the presence of larger CNVs.** We applied filters to remove outlier samples based on copy number estimates of the VNTR flanks: for each flanking region, we calculated the mean and StDev based on samples between the 30^th^ and 70^th^ percentiles of the population, defining outlier samples as those that were >7 StDevs from the mean and with consistent directionality for both flanks. Shown are two loci located within regions of known common copy number variation.^43^ For each locus, the top plot shows read depth of the 5’ flank, the middle plot shows read depth within the VNTR, and the bottom plot shows read depth within the 3’ flank. Samples in each plot are sorted based on read depth of the 3’ flank, with those that meet the criteria for being a consistent outlier for both flanks shown in red. Genotypes for these samples were not considered in downstream association analysis.

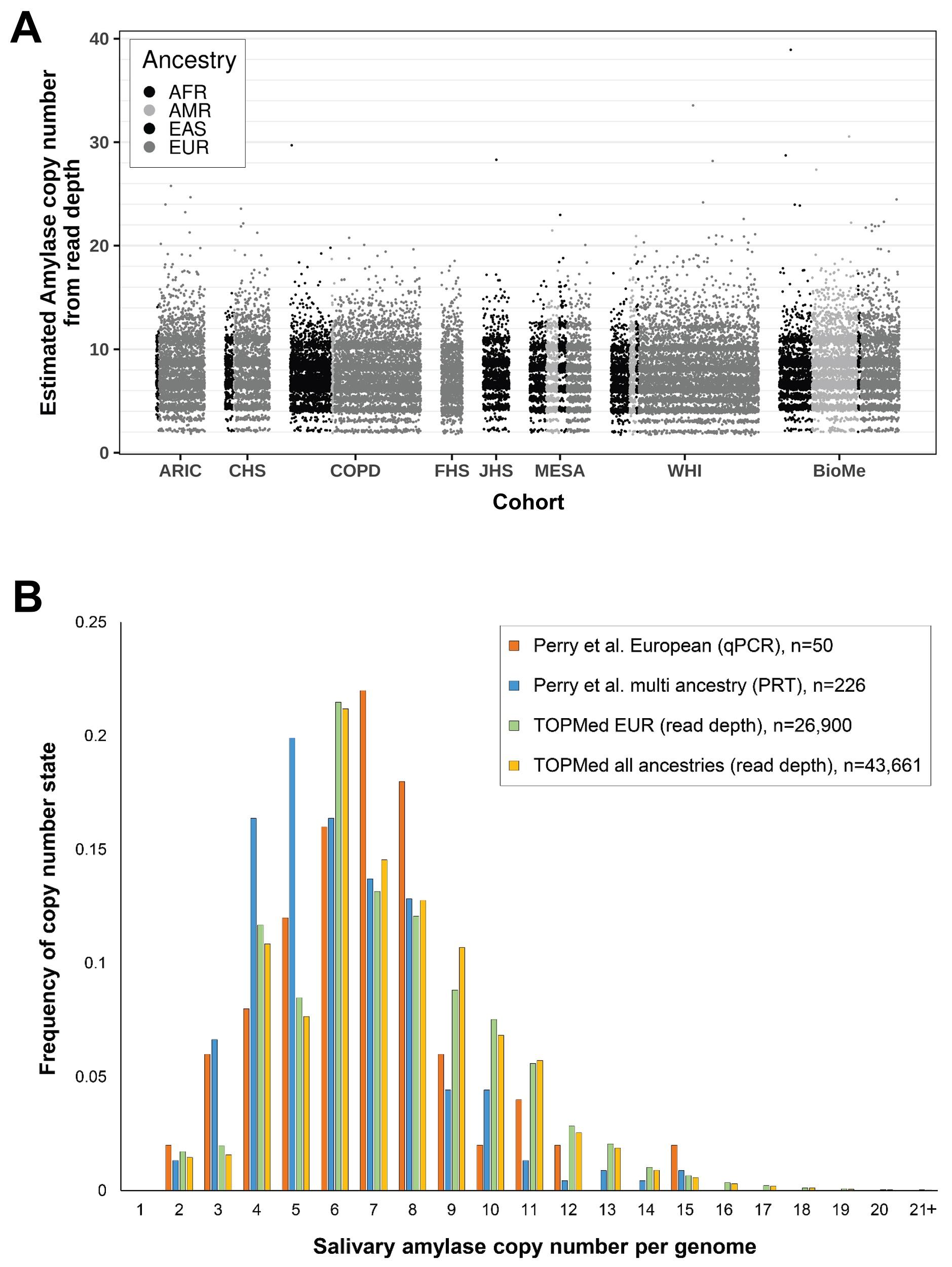

**Figure S2. Copy number estimates for salivary amylase genes. (A)** Absolute diploid copy number estimates generated using *mosdepth* for the salivary amylase 1 (*AMY1*) gene cluster at 1p21.1 in ~45,000 individuals from eight TOPMed cohorts used in this study. While most individuals carry between 2-15 copies of this locus, we observed rare individuals carrying up to an estimated 39 copies of *AMY1* genes. **(B)** Comparison of estimated copy numbers for the *AMY1* gene cluster obtained in TOPMed samples using read depth to those obtained in previously published cohorts using qPCR. The plot shows absolute copy number estimates for (i) European and multi-ancestry cohorts generated with qPCR published by Perry *et al.*,^44^ and (ii) TOPMed cohorts generated by *mosdepth* for the grouped 1p21.1 *AMY1* genes. In all cases, we present copy estimates rounded to the nearest integer. Both methods show similar frequency distributions, suggesting that the use of read depth yields accurate results.

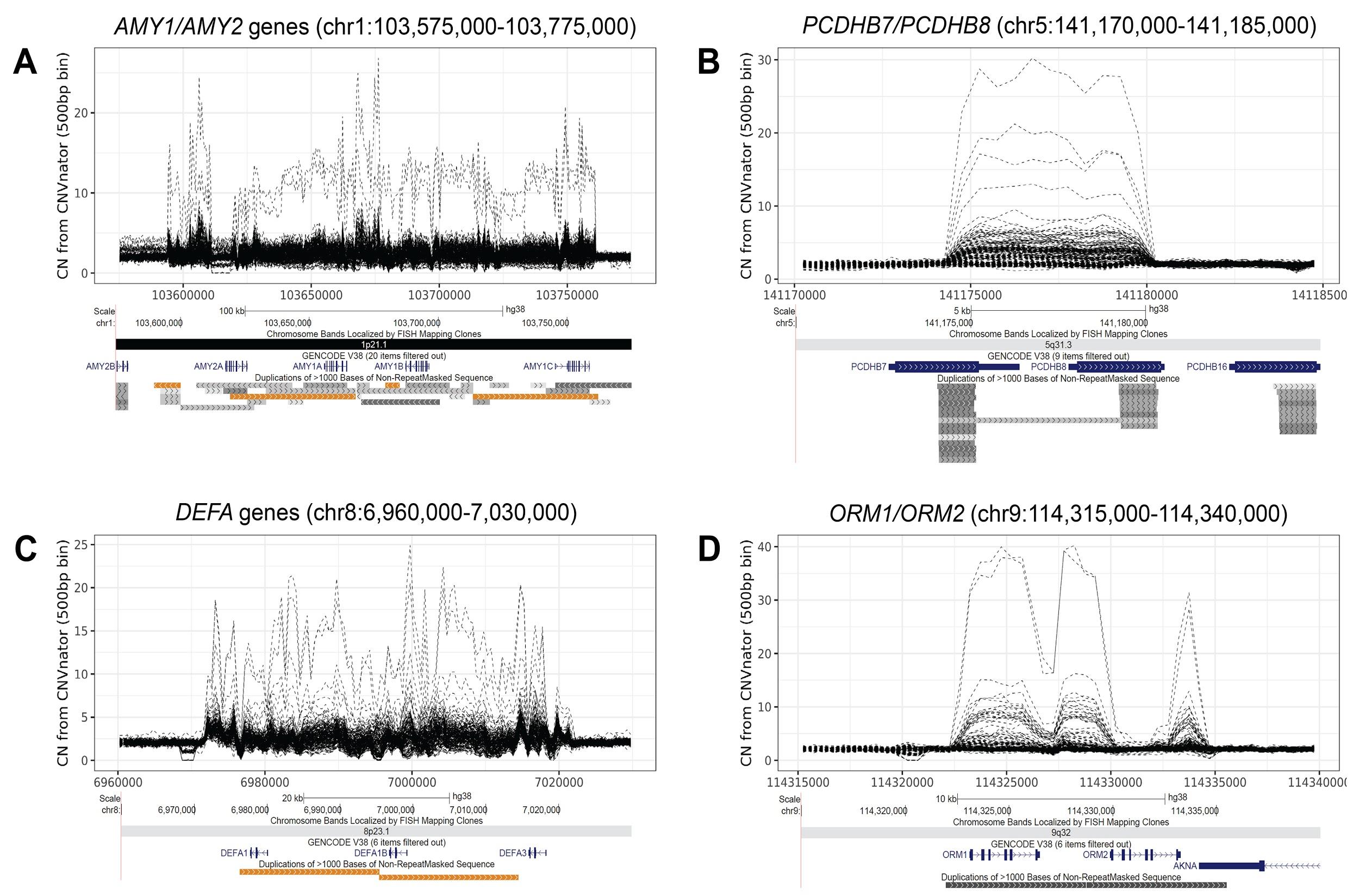

**Figure S3. Additional examples of genes showing extreme variation in copy number.** Using *mosdepth*, we generated copy number estimates for 1,105 multicopy genes in ~45,000 individuals. Within this cohort, we observed some genes that exhibited extreme variations in copy number, with some individuals having estimated copy numbers 10-20 times greater than the population average. To characterize these variants in more detail, we performed *CNVnator* analysis on 225 samples of interest, and plotted the estimated copy number across each locus. Shown are example plots of regions containing **(A)** *AMY1/AMY2* genes (chr1:103,575,000-103,775,000), **(B)** *PCDHB7/PCDHB8* (chr5:141,170,000-141,185,000), **(C)** *DEFA* genes (chr8:6,960,000-7,030,000), **(D)** *ORM1/ORM2* (chr9:114,315,000-114,340,000). Each plot shows *CNVnator* estimated relative diploid copy number per 500 bp bin in 225 individuals, with the copy number profile of each individual shown as a dashed line. Below each plot is an image of the region taken from the UCSC Genome Browser showing gene and segmental duplication annotations.

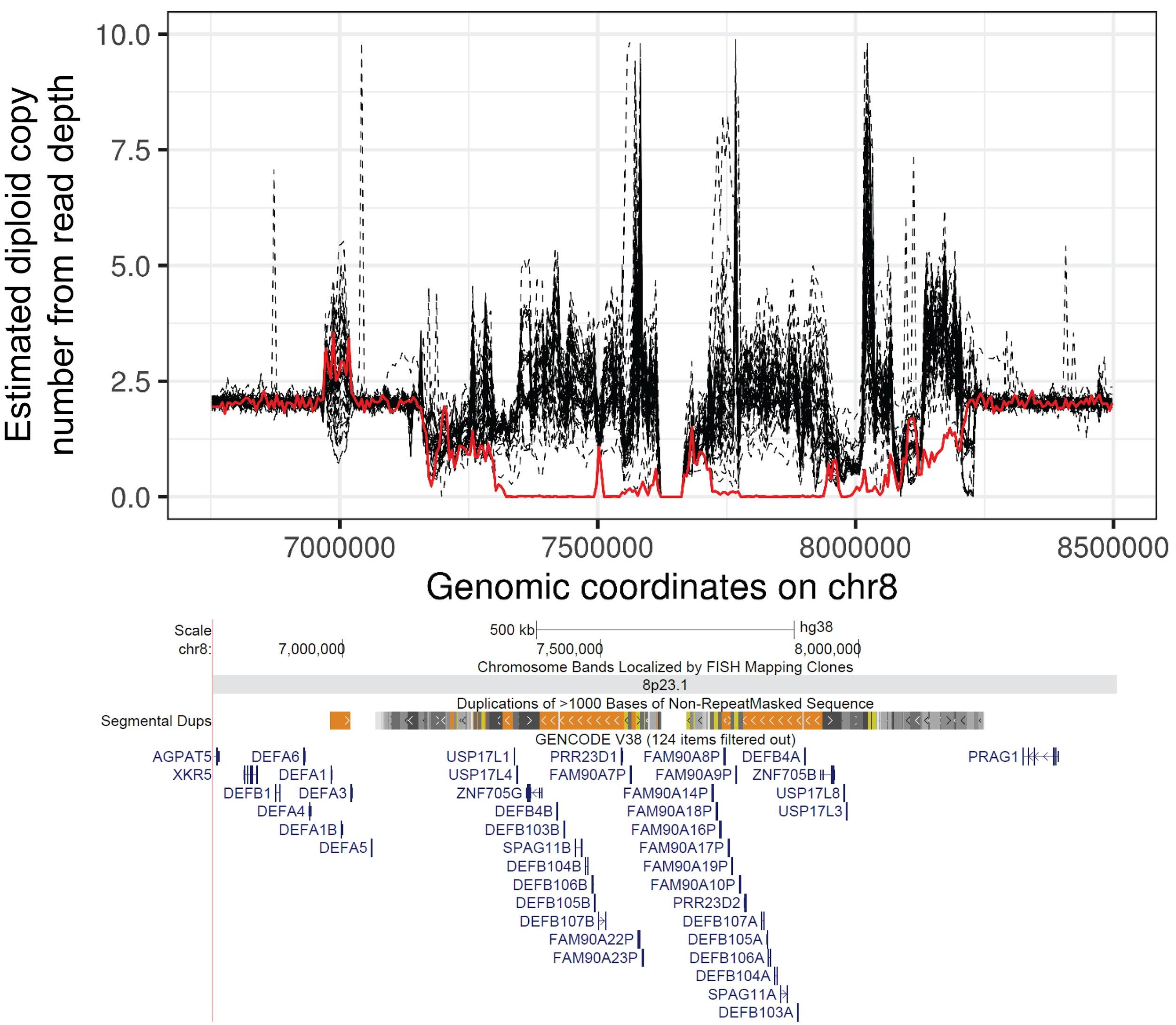

**Figure S4. Identification of individual with zero copies of the entire ꞵ-defensin gene cluster at 8p23.1**. Plot shows diploid copy number per 5 kb bin from *CNVnator* in 50 individuals for the ꞵ-defensin locus (chr8:6,750,001-8,500,000). Each line represents the copy number profile of one individual. The individual shown with the red line was originally identified using *mosdepth* as carrying ~zero copies of ꞵ-defensin genes in the region. Below the plot is an image of the region taken from the UCSC Genome Browser showing segmental duplication and gene annotations.

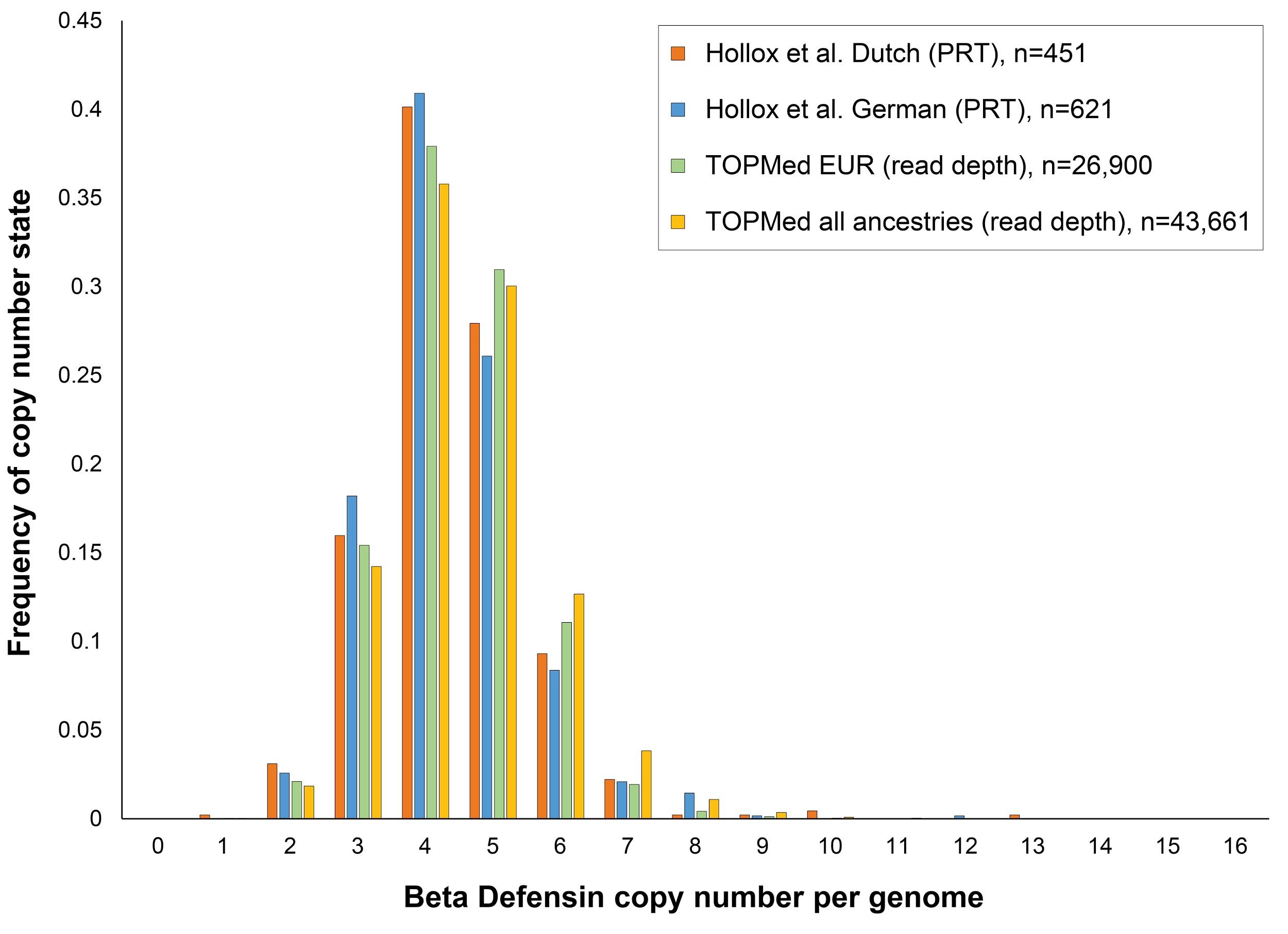

**Figure S5. Comparison of estimated copy numbers for the ꞵ-defensin gene cluster at 8p23.1 obtained in TOPMed samples using read depth to those obtained in previously published cohorts using the paralog ratio test.** The plot shows absolute copy number estimates for (i) two European cohorts generated with the paralog ratio test published by Hollox *et al.*,^7^ which is considered to be an accurate experimental method for quantifying multiallelic CNVs, and (ii) TOPMed cohorts generated by *mosdepth* for the grouped 8p23.1 ꞵ-defensin genes. In all cases, we present copy estimates rounded to the nearest integer. Both methods show highly similar frequency distributions, suggesting that the use of read depth yields accurate results.

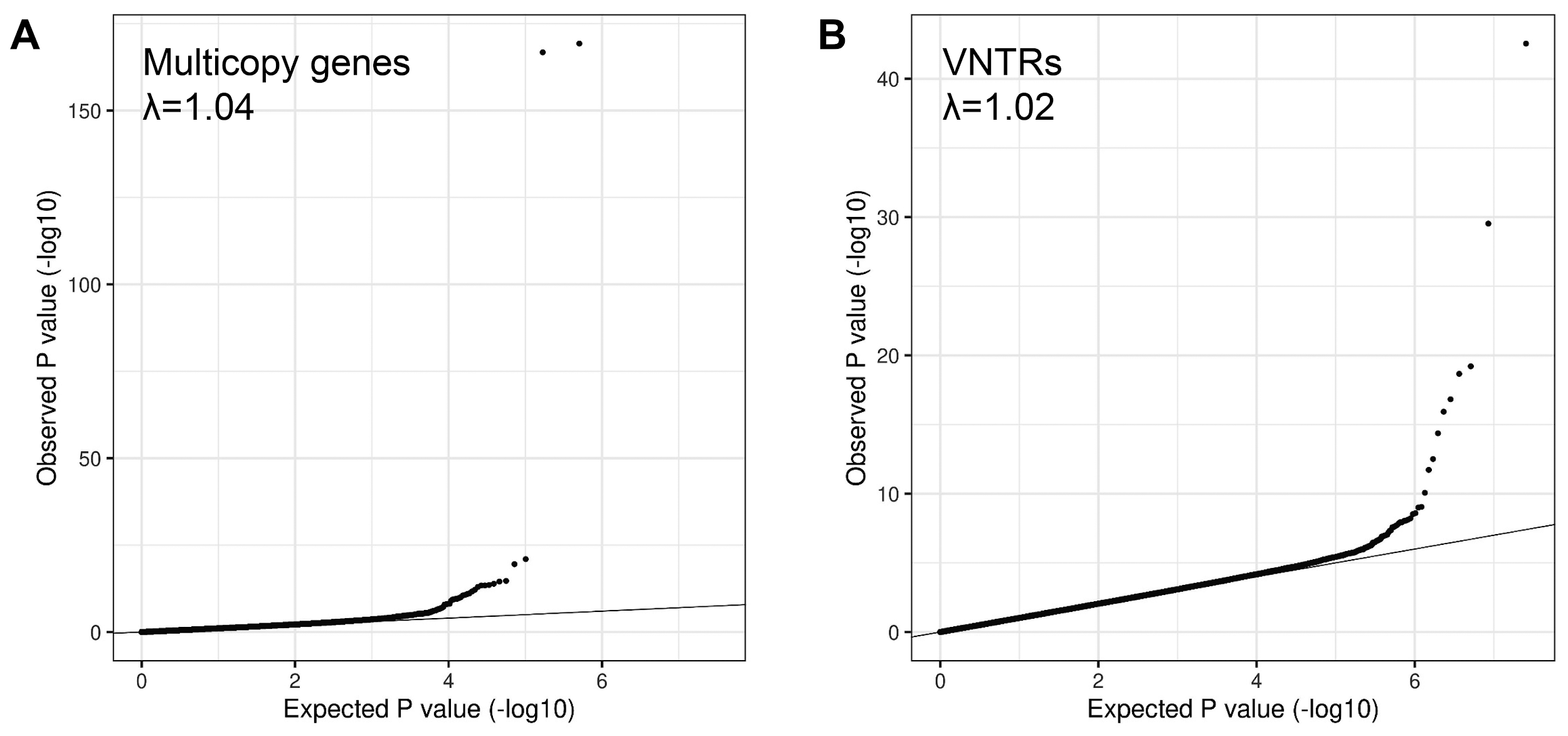

**Figure S6. QQ plots of meta-analysis discovery PheWAS using multicopy genes and VNTRs.** Genomic inflation was well controlled, with λ values between 1.00 and 1.04 for all ancestries tested.

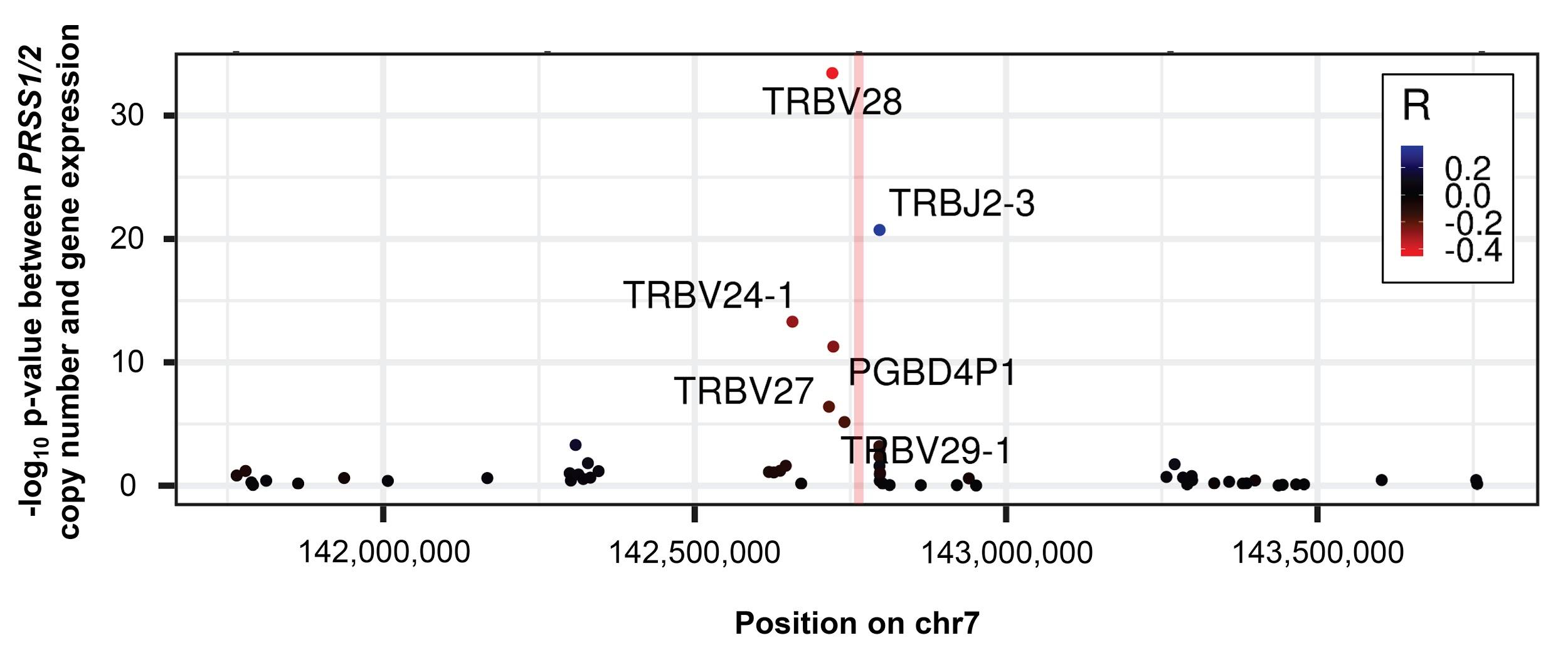

**Figure S7. Copy number of *PRSS1*/*PRSS2* correlates with the expression level of multiple neighboring T cell receptor ꞵ genes *in cis*.** Using eQTL analysis in the PPMI cohort, we observed that the expression level of multiple neighboring *TRB* genes showed significant correlations (both positive and negative) with copy number of *PRSS1/PRSS2*. The vertical red bar indicates the position of *PRSS1*/*PRSS2*, with each dot representing the -log_10_ p-value of association between estimated copy number of *PRSS1*/*PRSS2* and gene expression level from RNAseq in whole blood in the PPMI cohort. Points are colored based on the correlation value (R).

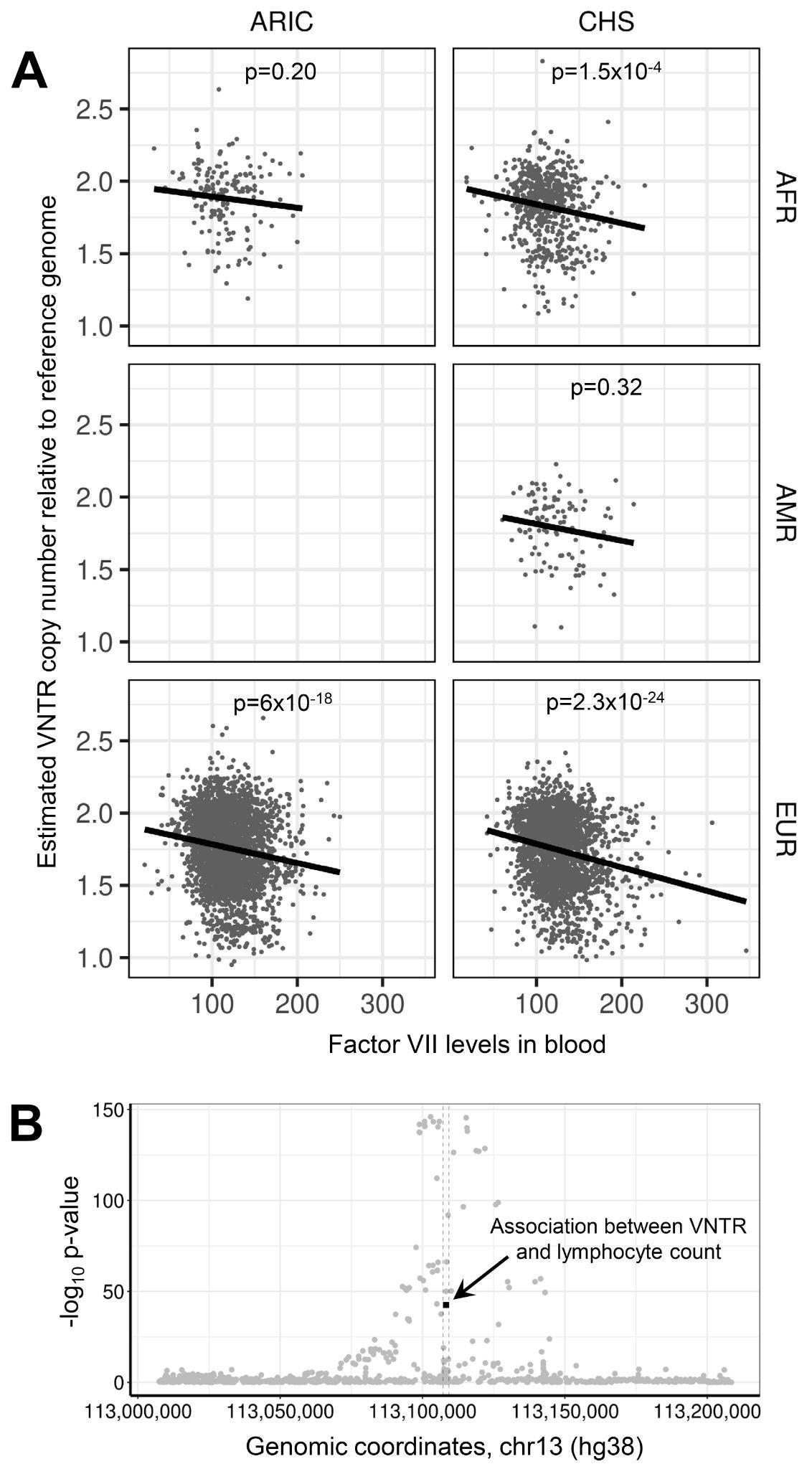

**Figure S8. Copy number of a 34mer tandem motif located within intron 1 of the *F7* gene at 13q34 (chr13:113,107,242-113,109,277) is not the causal variant associated with Factor VII levels in blood. (A)** We identified a strong and consistent association between copy number of this VNTR and Factor VII levels in blood across multiple TOPMed cohorts (discovery meta-analysis p=2.85x10^-43^). **(B)** We repeated the association analysis with Factor VII levels using all SNVs located within ±100 kb of the VNTR, which identified dozens of significant associations with local SNVs (grey circles), many of which showed much stronger associations than the one observed for VNTR copy number (black square). Using *MsCAVIAR*, we confirmed that VNTR copy number was not the likely causal variant to explain the observed association with Factor VII levels, with 24 SNVs ranked by *MsCAVIAR* as having higher probabilities of being causal compared to the VNTR (Table S8).

**Acknowledgements**

This work was supported by NIH grants NS105781 and NS120241 to AJS, and NIH predoctoral fellowship NS108797 to OR. Research reported in this paper was supported by the Office of Research Infrastructure of the National Institutes of Health under award number S10OD018522. The content is solely the responsibility of the authors and does not necessarily represent the official views of the National Institutes of Health. This work was supported in part through the computational resources and staff expertise provided by Scientific Computing at the Icahn School of Medicine at Mount Sinai.

Molecular data for the Trans-Omics in Precision Medicine (TOPMed) program was supported by the National Heart, Lung and Blood Institute (NHLBI). Genome sequencing for “NHLBI TOPMed - NHGRI CCDG: The BioMe Biobank at Mount Sinai” (phs001644.v1.p1) was performed at the McDonnell Genome Institute (3UM1HG008853-01S2). Genome sequencing for “NHLBI TOPMed: Women's Health Initiative (WHI)” (phs001237.v2.p1) was performed at the Broad Institute Genomics Platform (HHSN268201500014C). Core support including centralized genomic read mapping and genotype calling, along with variant quality metrics and filtering were provided by the TOPMed Informatics Research Center (3R01HL-117626-02S1; contract HHSN268201800002I). Core support including phenotype harmonization, data management, sample-identity QC, and general program coordination were provided by the TOPMed Data Coordinating Center (R01HL-120393; U01HL-120393; contract HHSN268201800001I). We gratefully acknowledge the studies and participants who provided biological samples and data for TOPMed.

The Women’s Health Initiative (WHI) program is funded by the National Heart, Lung, and Blood Institute, National Institutes of Health, U.S. Department of Health and Human Services through contracts HHSN268201600018C, HHSN268201600001C, HHSN268201600002C, HHSN268201600003C, and HHSN268201600004C. This manuscript was not prepared in collaboration with investigators of the WHI, and does not necessarily reflect the opinions or views of the WHI investigators, or NHLBI.

The Atherosclerosis Risk in Communities study has been funded in whole or in part with Federal funds from the National Heart, Lung, and Blood Institute, National Institute of Health, Department of Health and Human Services, under contract numbers (HHSN268201700001I, HHSN268201700002I, HHSN268201700003I, HHSN268201700004I, and HHSN268201700005I). The authors thank the staff and participants of the ARIC study for their important contributions

MESA and the MESA SHARe project are conducted and supported by the National Heart, Lung, and Blood Institute (NHLBI) in collaboration with MESA investigators. Support for MESA is provided by contracts

HHSN268201500003I, N01-HC-95159, N01-HC-95160, N01-HC-95161, N01-HC-95162, N01-HC-95163, N01-HC95164, N01-HC-95165, N01-HC-95166, N01-HC-95167, N01-HC-95168, N01-HC-95169, UL1-TR-001079, UL1-TR000040, UL1-TR-001420, UL1-TR-001881, and DK063491.

The Jackson Heart Study (JHS) is supported and conducted in collaboration with Jackson State University (HHSN268201800013I), Tougaloo College (HHSN268201800014I), the Mississippi State Department of Health (HHSN268201800015I/HHSN26800001) and the University of Mississippi Medical Center (HHSN268201800010I, HHSN268201800011I and HHSN268201800012I) contracts from the National Heart, Lung, and Blood Institute (NHLBI) and the National Institute for Minority Health and Health Disparities (NIMHD). The authors also wish to thank the staff and participants of the JHS.

This research was supported by contracts HHSN268201200036C, HHSN268200800007C, HHSN268201800001C, N01HC55222, N01HC85079, N01HC85080, N01HC85081, N01HC85082, N01HC85083, N01HC85086, and grants U01HL080295 and U01HL130114 from the National Heart, Lung, and Blood Institute (NHLBI), with additional contribution from the National Institute of Neurological Disorders and Stroke (NINDS). Additional support was provided by R01AG023629 from the National Institute on Aging (NIA). A full list of principal CHS investigators and institutions can be found at CHS-NHLBI.org.

This research used data generated by the COPDGene study, which was supported by NIH grants U01 HL089856 and U01 HL089897. The COPDGene project is also supported by the COPD Foundation through

contributions made by an Industry Advisory Board comprised of Pfizer, AstraZeneca, Boehringer Ingelheim,

Novartis, and Sunovion.

The Framingham Heart Study is conducted and supported by the National Heart, Lung, and Blood Institute (NHLBI) in collaboration with Boston University (Contract No. N01-HC-25195, HHSN268201500001I and 75N92019D00031). This manuscript was not prepared in collaboration with investigators of the Framingham Heart Study and does not necessarily reflect the opinions or views of the Framingham Heart Study, Boston University, or NHLBI.

The Mount Sinai BioMe Biobank is supported by The Andrea and Charles Bronfman Philanthropies.	Data used in the preparation of this article were obtained from the Parkinson’s Progression Markers Initiative (PPMI) database (www.ppmiinfo.org/data). For up-to-date information on the study, visit [www.ppmiinfo.org](http://www.ppmiinfo.org/). PPMI, a public-private partnership, is funded by the Michael J. Fox Foundation for Parkinson’s Research and funding partners, a full list of which can be found at [www.ppmiinfo.org/fundingpartners](http://www.ppmiinfo.org/fundingpartners).

The Genotype-Tissue Expression (GTEx) Project was supported by the Common Fund of the Office of the Director of the National Institutes of Health (commonfund.nih.gov/GTEx). Additional funds were provided by the NCI, NHGRI, NHLBI, NIDA, NIMH, and NINDS. Donors were enrolled at Biospecimen Source Sites funded by NCI\Leidos Biomedical Research, Inc. subcontracts to the National Disease Research Interchange (10XS170), Roswell Park Cancer Institute (10XS171), and Science Care, Inc. (X10S172). The Laboratory, Data Analysis, and Coordinating Center (LDACC) was funded through a contract (HHSN268201000029C) to The Broad Institute, Inc. Biorepository operations were funded through a Leidos Biomedical Research, Inc. subcontract to Van Andel Research Institute (10ST1035). Additional data repository and project management were provided by Leidos Biomedical Research, Inc.(HHSN261200800001E). The Brain Bank was supported by supplements to University of Miami grant DA006227. Statistical Methods development grants were made to the University of Geneva (MH090941 & MH101814), the University of Chicago (MH090951, MH090937, MH101825, & MH101820), the University of North Carolina - Chapel Hill (MH090936), North Carolina State University (MH101819), Harvard University (MH090948), Stanford University (MH101782), Washington University (MH101810), and to the University of Pennsylvania (MH101822). The datasets used for the analyses described in this manuscript were obtained from dbGaP at http://www.ncbi.nlm.nih.gov/gap through dbGaP accession number phs000424.v7.p2.
